# Vaccinating children against COVID-19: commentary and mathematical modelling

**DOI:** 10.1101/2022.01.05.22268820

**Authors:** Michael T. Hawkes, Michael F. Good

**Author notes:** Address correspondence to Michael T. Hawkes,; and Michael F Good,.

## Abstract

With the recent licensure of mRNA vaccines against COVID-19 in the 5-11 year old age group, the public health impact of a childhood immunization campaign is of interest. Using a mathematical epidemiological model, we project that childhood vaccination carries minimal risk and yields modest public health benefits. These include large relative reductions in child morbidity and mortality, although the absolute reduction is small because these events are rare. Furthermore, the model predicts “altruistic” absolute reductions in adult cases, hospitalizations, and mortality. However, vaccinating children to benefit adults should be considered from an ethical as well as a public health perspective. From a global health perspective, an additional ethical consideration is the justice of giving priority to children in high-income settings at low risk of severe disease while vaccines have not been made available to vulnerable adults in low-income settings.

## Introduction

Recently, Pfizer and BioNTech provided evidence that their mRNA vaccine against SARS-CoV-2 is safe and immunogenic in children 5 to 11 years of age.^1^ As countries prepare to incorporate school-age children into their vaccination schedules, the public health risks and benefits, as well as ethical considerations should be carefully weighed.

One of the most distinguishing features of the SARS-CoV-2 pandemic is the dramatic correlation between age and disease severity. Early in the pandemic, a study in Switzerland from February to June 2020, demonstrated that the infection fatality rate for those over 65 years was 5.6 per 100.^2^ This compared with a rate of 0.0016 per 100 in those aged 5-9 years and only 0.00032 per 100 for those aged 10-19 years.^2^ Data from the USA show that over 19 months, there were 349 deaths in those aged 0-17 years from a total of 606,389 deaths.^3^ Based on these figures, the chances of a child (<18 years old) dying from COVID-19 was ∼ 2,500 times less than that of an older American (>65 years old). Clearly, early on in the pandemic, although children were dying of COVID-19, it was at a very low rate. On the other hand, the emergence of the more infectious SARS-CoV-2 delta variant has led to increased cases in children and there is increased recognition of paediatric morbidity associated with the multisystem inflammatory syndrome in children (MIS-C). Nonetheless, based on these figures, the benefits of vaccination are far greater for the elderly than for children, but there is a dynamic interplay of benefits and risks that needs to be considered.

Vaccines against SARS-CoV-2 are highly effective in clinical trials and in real-world settings.^4^ Pfizer and BioNTech recently announced that their mRNA vaccine is safe and generates robust neutralizing antibody levels in children 5 to 11 years of age.^1^

The public health benefits of vaccination are twofold: to protect the health of vaccinees and to contribute to herd immunity. Herd immunity occurs when the percentage of the population who are unable to transmit the virus as a result of immunity is sufficient to extinguish the epidemic. *ℛ*_0_ is the basic reproduction number for a virus in a non-immune population. Immunity in the population occurs as a result of vaccination and natural immunity (as a result of infection). Using the number of ‘cases’ as a proxy for the number of infections and hence the number with natural immunity, in the USA and the UK approximately 10% of the population have natural immunity whereas in Australia, 0.16% have natural immunity. However, these numbers are underestimates as many people who have been infected remain asymptomatic and do not become ‘cases’.^5^

If vaccinated individuals cannot transmit the virus, then the fraction of the population that needs to be vaccinated to end the epidemic is approximately 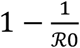. The *ℛ*_0_ for the dominant delta strain of SARS-CoV-2 has been reported to be between 5 and 8.^6^ This means that between 80% and 87.5% of the entire population need to be immune and non-transmitting. Even if ∼10% of the population were to develop immunity as a result of natural infection, then still between 70-80% of the population will need to be vaccinated and non-transmitting. There will be a need to vaccinate children of all ages, as well as adults, to reach 70-80%. Furthermore, recent reports show that the risk of household transmission is reduced by only ∼50% among vaccinated people who develop breakthrough SARS-CoV-2 infection.^7^

While herd immunity is unlikely to be attainable through vaccination, higher levels of immunity will reduce the spread. Combined with social distancing and wearing of masks, we will be able to control focal epidemics.^8^

To hasten and enhance the development of herd immunity, vaccination of children 5-11 years of age may be contemplated (∼10% of the entire population). However, vaccinating young children will have limited direct benefit to them as outlined above, but this population, by being immune will protect the older and more vulnerable, and in particular the non-vaccinated.

We hypothesize that childhood vaccination against SARS-CoV-2 will be associated with reductions in disease burden in children (directly, through disease attenuation among vaccinees) and in adults (indirectly, through herd immunity). We use mathematical modelling to support this hypothesis and provide quantitative estimates of the public health effects of childhood vaccination over one year in two jurisdictions: Australia and Alberta (Canada).

## Methods

### Description of the model and model parameters

We used a deterministic Susceptible-Infected-Recovered (SIR) compartmental model with age structure (seven strata: under 5 years of age; 5-11; 12-19; 20-39; 40-59; 60-74; 75 and older) and vaccine with imperfect efficacy. The flow chart for this model is shown in Figure 1. This model accounts for vital dynamics: births (*∧*); aging between strata (*α*_*i*_); and age-specific natural death rate (*μ*_*i*_). The model includes the following infection parameters: standard incidence ratio (*β*); contact rate based on published social contact matrices^9^ (*cm*_*ij*_); age-specific relative infectiousness (*τ*_*i*_); age-specific relative susceptibility to infection (*σ*_*i*_); duration of infection (1*/δ*); and age-specific infection fatality rate (*f*_*i*_). The effect of vaccination was modelled as relative reduction in: infectiousness (*ε*_*τ*_); susceptibility to acquiring infection (*ε*_*σ*_); probability of hospitalization (*ε*_*h*_); and mortality (*ε*_*f*_). To model the changes in the contact rate due to public health measures, we used a “social distancing” parameter, *θ*, which varied from zero (complete lockdown) to 1 (complete mixing in the population), as described in a previous modelling study.^8^ The mathematical properties of this model have been previously discussed.^10^

**Figure 1.**
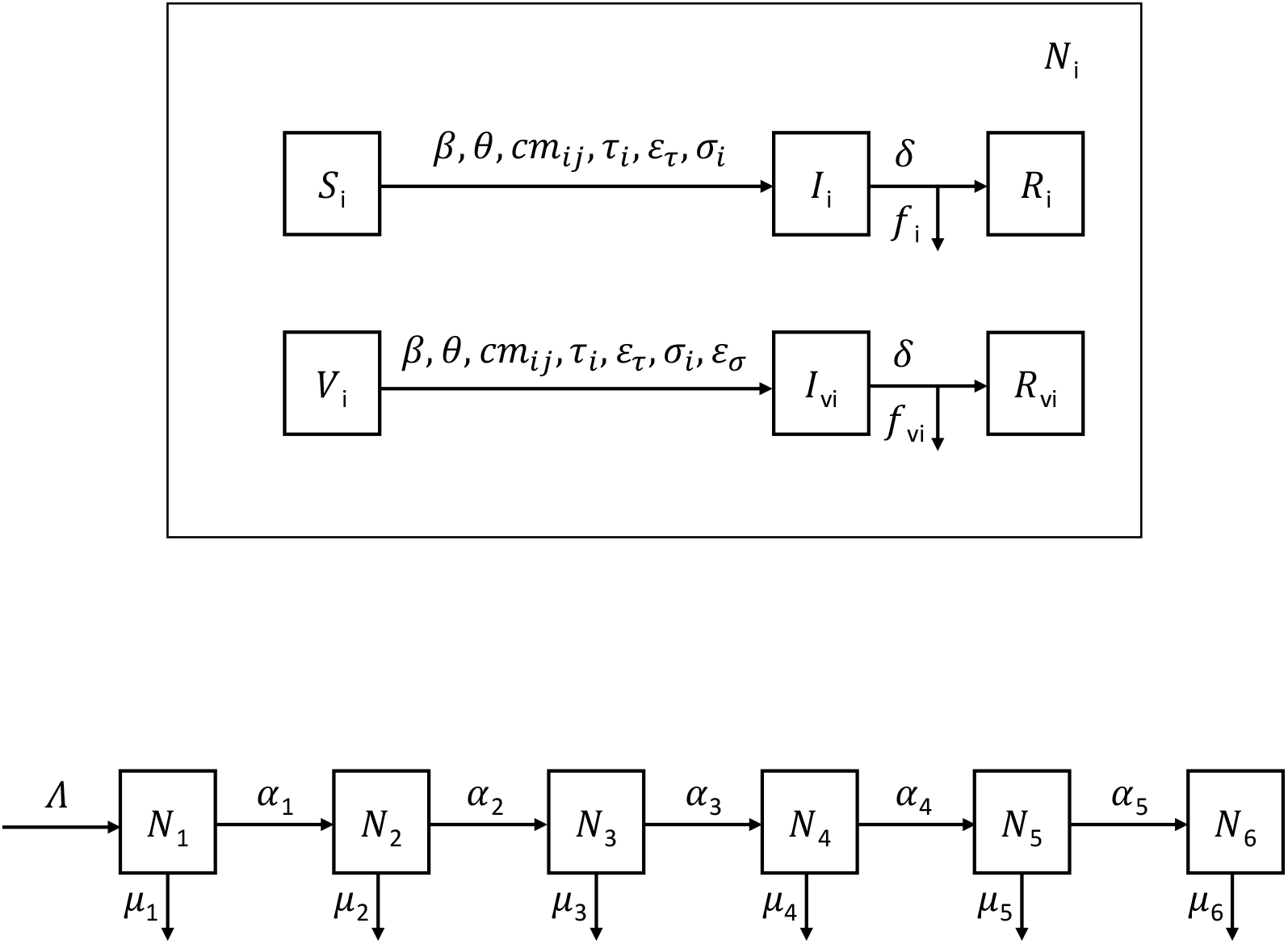
Flow chart for model. The Susceptible-Infected-Recovered (SIR) compartmental model was divided into 7 age classes. This allowed us to incorporate age-specific parameters in the model: birth rate (*Λ*), aging between strata (*α*_*i*_), natural mortality rates (*μ*_*i*_), social contact matrix (*cm*_*ij*_), relative infectiousness with SARS-CoV-2 (*τ*_*i*_), relative susceptibility (*σ*_*i*_), hospitalization (*h*_*i*_), and fatality rates (*f*_*i*_). Estimates for the transmission rate (*β*) and the duration of infection (1*/δ*) were taken from previous studies. A parameter theta (*θ*), reflecting the intensity of public health measures to prevent transmission (e.g., social distancing, mask mandates, service closures) was included to account for reduction in the contact rate from the assumption of perfect mixing. The effect of vaccination was modelled by four parameters: proportional reduction in infectiousness (*ε*_*τ*_), susceptibility (*ε*_*σ*_), hospitalization (*ε*_*h*_), and mortality (*ε*_*f*_).

Parameter estimates are shown in Table 1. We used realistic estimates based on vital statistics in Australia and Alberta, as well as biological characteristics of SARS-CoV-2. We modelled both the currently dominant and highly infectious delta variant (*ℛ*_0_ =5.08) and the historically important alpha variant (*ℛ*_0_ =2.79).

**Table 1.**
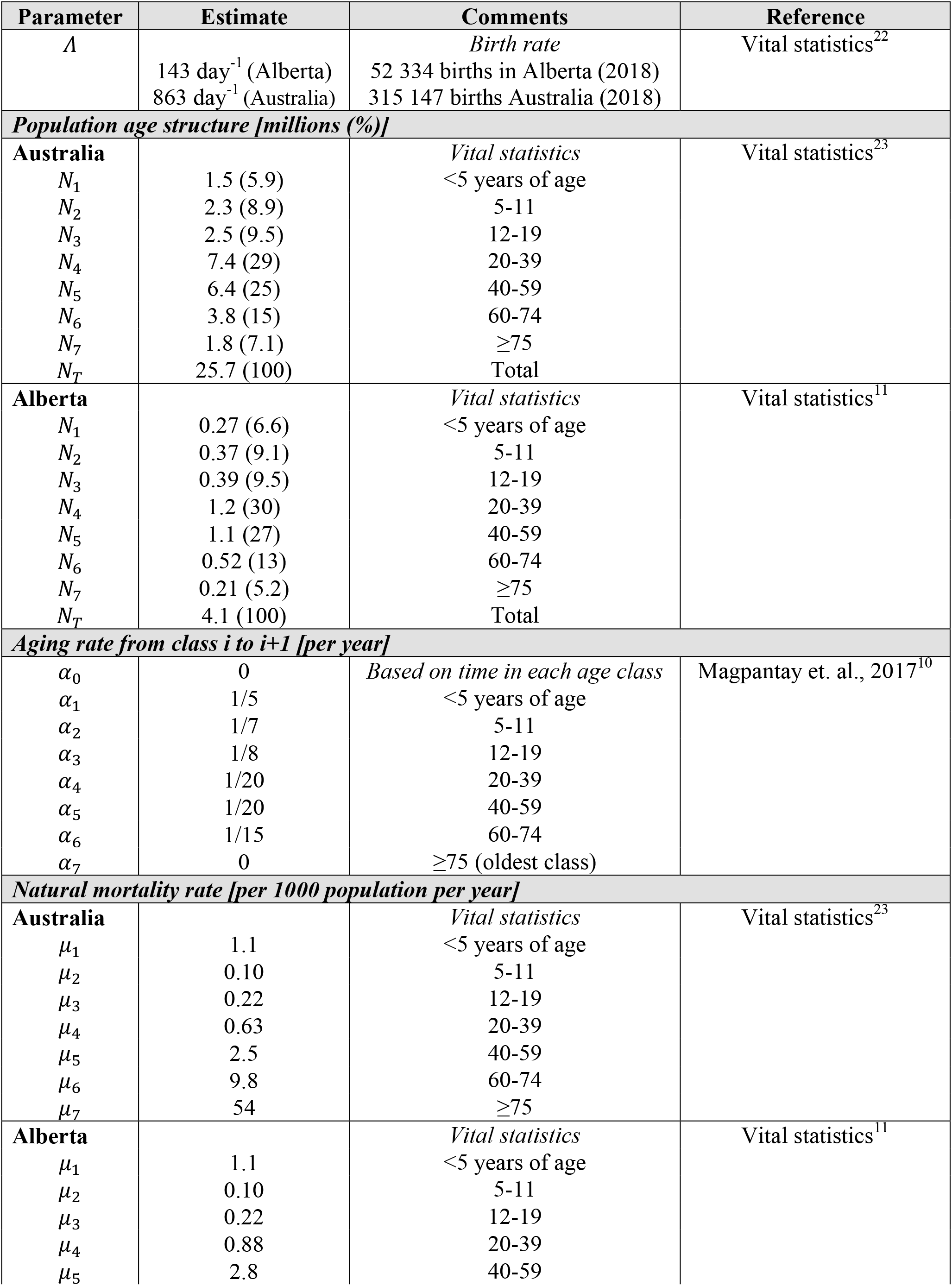
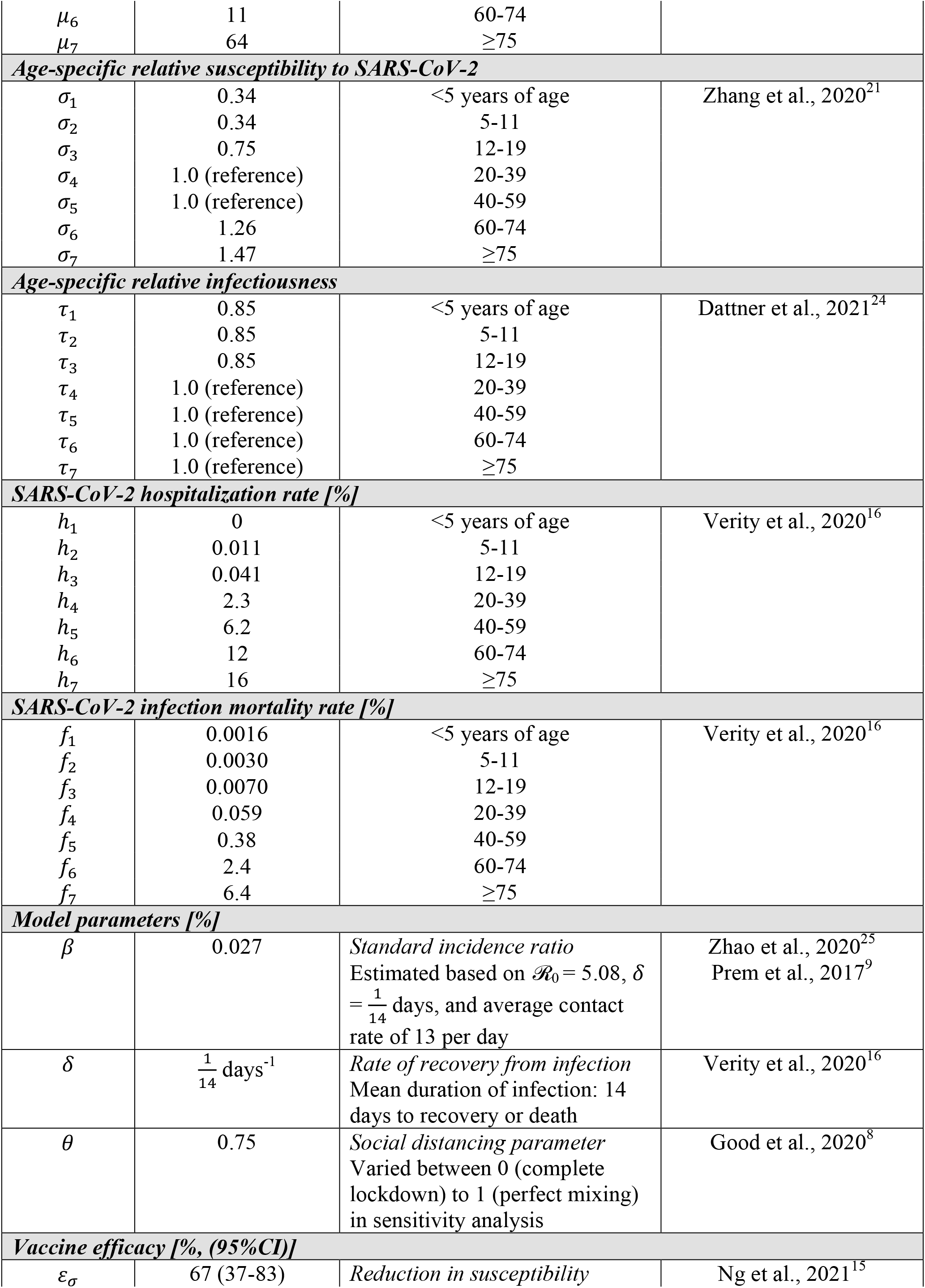
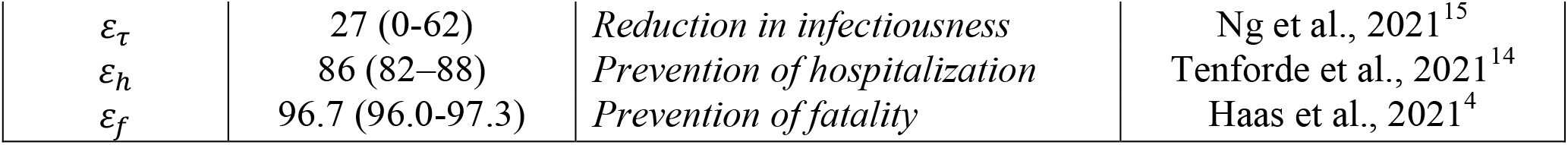
Model parameters: values and rationale.

A system of 49 ordinary differential equations (ODEs) describes the flow between compartments:

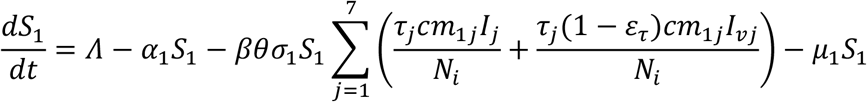

For *i* =2,…,7

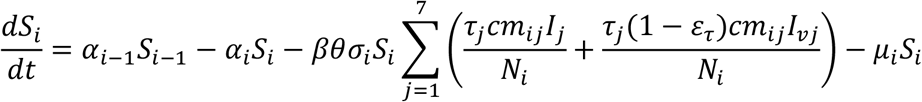

For *i* =1,2…,7

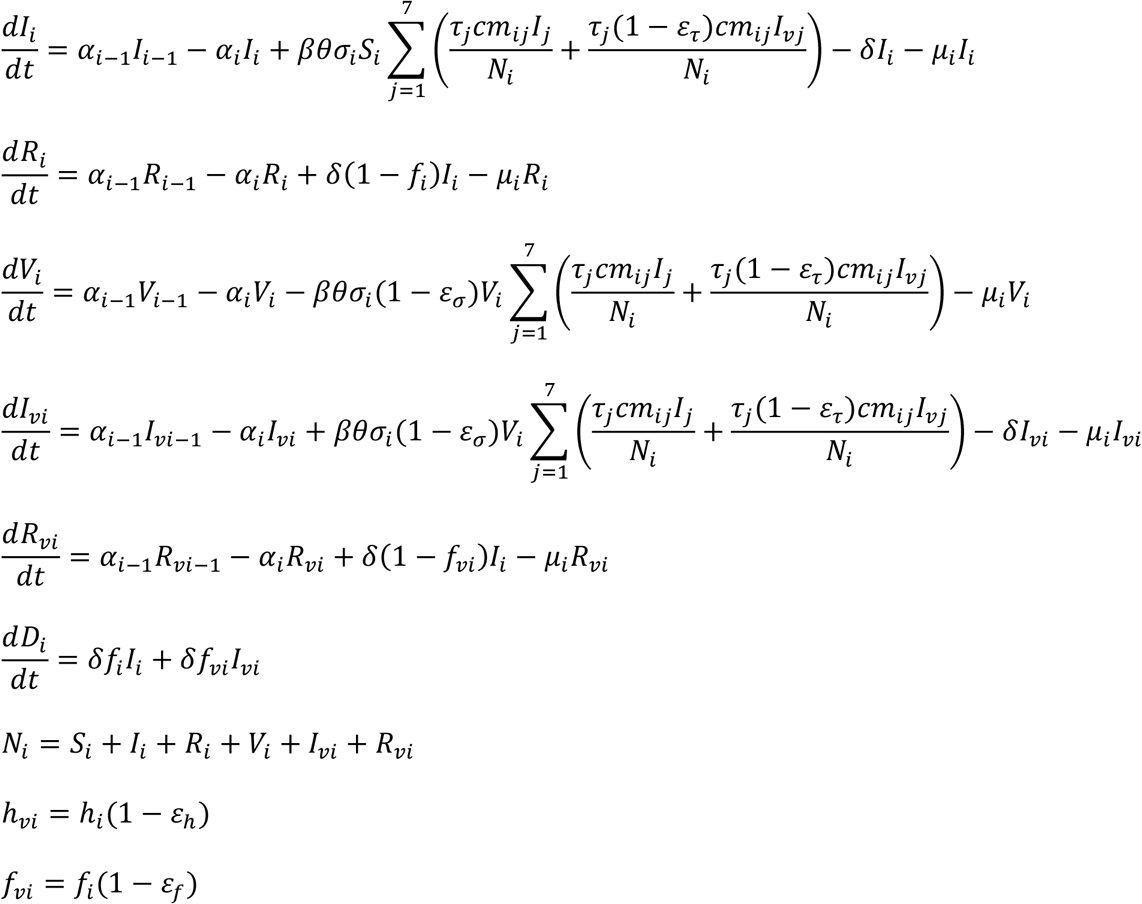

### Initial conditions and time horizon

To model the course of a future “wave” of SARS-CoV-2, we began with initial conditions which included the total population of Australia or Alberta, divided into age classes, and further subdivided into vaccinated and unvaccinated compartments.^11^ The proportion of actively infected individuals on was calculated based on the number of known active cases in Australia or Alberta in August 2021, proportionally divided among the age classes. We assumed that all these cases would be isolated, and therefore would not contribute to the infectious pool. We further assumed that a five-fold higher number of undiagnosed cases would be present in the community. This assumption is based on a previous study, which estimated that the number of infections in the US was 3 to 20 times higher than the number of confirmed cases.^5^ The proportion of recovered individuals was based on seroprevalence surveys.^12,13^ In hypothetical scenarios for Australia, 80% or 90% of adults were presumed to be vaccinated at baseline. In the Alberta scenario, the proportion of vaccinated individuals in each age stratum in October 2021 was used as the baseline. The model was run with no childhood vaccination and with 80% childhood vaccination. The model was run for a period of 365 days. In the absence of an analytical solution to the system of ODEs, we used numerical simulations (package *deSolver*) in the *R* statistical environment (R version 3.6.2).

### Confidence intervals for model outputs

To account for uncertainties in the vaccine efficacy, we used a multi-way sensitivity analysis, varying the four key parameters (efficacy to prevent transmission, susceptibility, hospitalization, and death) over their 95% confidence interval, based on published studies.^4,14,15^ We assumed that each proportion followed a beta-distribution. We randomly sampled from the distribution of each parameter, used these as inputs for the model, and ran the SIR model 1000 times. Using the distribution of model outputs that was generated, the 95% confidence interval for each output was defined by the 2.5^th^ percentile and the 97.5^th^ percentile.

### Variation of model estimates with vaccine uptake and concurrent public health measures

The proportion of vaccinated children and public health measures may vary widely over time and geography. Therefore, we varied these key parameters from 0 to 1 (over the entire possible range) and plotted the resulting model outputs. Graphical methods were used to examine the dependence of model outputs on key parameters.

## Results

We modelled the course of SARS-CoV-2 delta variant (*ℛ*_0_=5.08) in Australia (Table 2) and Alberta (Table 3) for one year, with and without childhood vaccination. The expected epidemic curve was observed, with infections eventually extinguishing to zero (Figure 2).

**Table 2.** Simulation for Australia (*ℛ*_0_ = 5.08, 80% of adults vaccinated): Projected differences in cases, hospitalizations, deaths due to COVID-19, multisystem inflammatory syndrome in children (MIS-C) and vaccine adverse events associated with childhood vaccination.

|  | No childhood vaccination | Childhood vaccination<br>(80% coverage) | Absolute reduction | Relative reduction<br>(%) |
| --- | --- | --- | --- | --- |
| **Cases of COVID-19 (×1000)** |  |  |  |  |
| All age groups | 12200 (3790-18200) | 10500 (1610-17400) | 1760 (845-2560) | 14 (4.6-59) |
| 5 to 11 years old | 729 (262-1030) | 233 (25.9-564) | 496 (241-563) | 68 (45-91) |
| Vaccinated adults | 7800 (1440-13000) | 6860 (635-12700) | 932 (258-1440) | 12 (2-58) |
| Unvaccinated adults | 3530 (1820-3950) | 3220 (850-3850) | 305 (104-993) | 8.6 (2.6-54) |
| **Hospitalizations<sup>1</sup>** |  |  |  |  |
| All age groups | 532000 (152000-857000) | 472000 (68800-834000) | 60600 (23200-97700) | 11 (2.7-56) |
| 5 to 11 years old | 78.4 (28.2-110) | 25.1 (2.78-60.6) | 53.3 (25.9-60.5) | 68 (45-91) |
| Vaccinated adults | 360000 (62700-650000) | 314000 (27700-630000) | 46500 (17700-65800) | 13 (2.9-58) |
| Unvaccinated adults | 172000 (79000-208000) | 158000 (37700-204000) | 14000 (3860-42500) | 8.2 (1.9-53) |
| **Deaths<sup>1</sup>** |  |  |  |  |
| All age groups | 30100 (12900-39000) | 27200 (6230-38000) | 2870 (1020-6740) | 9.6 (2.6-52) |
| 5 to 11 years old | 22.0 (7.79-31.0) | 3.53 (0.589-6.24) | 18.5 (7.19-24.8) | 84 (80-93) |
| Vaccinated adults | 1830 (289-3680) | 1590 (133-3560) | 241 (103-333) | 13 (3.3-58) |
| Unvaccinated adults | 28200 (12300-35600) | 25600 (6060-34800) | 2610 (893-6500) | 9.3 (2.5-51) |
| **MIS-C cases (0-19 years old)** | 230 (82.9-325) | 73.7 (8.19-178) | 157 (76.1-178) | 68 (45-91) |
| **Vaccine-related adverse events** |  |  |  |  |
| Myocarditis | 20 (9.0-123) | 38 (18-237) | -18 (-3 to -112)<sup>2</sup> | -93 (-15 to -570)<sup>2</sup> |
| Anaphylaxis | 22 (9.8-35) | 42 (19-68) | -20 (-8.1 to -50)<sup>2</sup> | -92 (-37 to -230)<sup>2</sup> |

<sup>1</sup>Due to acute COVID-19
<sup>2</sup>Negative sign indicates increase in cases with vaccination

**Table 3.** Simulation for Alberta (*ℛ*_0_ = 5.08): Projected differences in cases, hospitalizations, deaths due to COVID-19, multisystem inflammatory syndrome in children (MIS-C) and vaccine adverse events associated with childhood vaccination.

|  | **No childhood vaccination** | **Childhood vaccination (80% coverage)** | **Absolute reduction** | **Relative reduction (%)** |
| --- | --- | --- | --- | --- |
| **Cases of COVID-19 (×1000)** |  |  |  |  |
| All age groups | 1950 (788-2870) | 1750 (595-2760) | 206 (114-237) | 11 (4-25) |
| 5 to 11 years old | 97.3 (45.8-140) | 34.6 (9.16-78.1) | 62.7 (36.7-72.7) | 64 (43-82) |
| Vaccinated adults | 1190 (284-1990) | 1090 (217-1950) | 104 (35.5-129) | 8.7 (1.9-24) |
| Unvaccinated adults | 608 (405-667) | 575 (327-654) | 33.3 (12.6-76.8) | 5.5 (1.9-19) |
| **Hospitalizations<sup>1</sup>** |  |  |  |  |
| All age groups | 76800 (27100-123000) | 70600 (21700-120000) | 6180 (2770-7900) | 8 (2.2-21) |
| 5 to 11 years old | 10.5 (4.92-15) | 3.72 (0.984-8.39) | 6.74 (3.94-7.81) | 64 (43-82) |
| Vaccinated adults | 55200 (12500-98800) | 50000 (9510-96000) | 5210 (2260-6210) | 9.4 (2.6-24) |
| Unvaccinated adults | 21600 (13800-24400) | 20600 (11300-24200) | 959 (247-2460) | 4.4 (1-18) |
| **Deaths<sup>1</sup>** |  |  |  |  |
| All age groups | 3870 (2540-4570) | 3700 (2160-4500) | 177 (63.4-376) | 4.6 (1.4-15) |
| 5 to 11 years old | 3.04 (1.48-4.32) | 0.61 (0.286-0.964) | 2.43 (1.2-3.36) | 80 (78-81) |
| Vaccinated adults | 259 (57-512) | 234 (42.7-498) | 25.2 (11.5-30.6) | 9.7 (2.9-24) |
| Unvaccinated adults | 3610 (2450-4090) | 3460 (2090-4040) | 149 (46.7-358) | 4.1 (1.1-15) |
| **MIS-C cases (0-19 years old)** | 30.7 (14.5-44.2) | 10.9 (2.89-24.7) | 19.8 (11.6-23) | 64 (43-82) |
| **Vaccine-related adverse events** |  |  |  |  |
| Myocarditis | 2.8 (1.3-18) | 5.8 (2.8-36) | -3.0 (-0.48 to -18)<sup>2</sup> | -105 (-17 to -650)<sup>2</sup> |
| Anaphylaxis | 3.1 (1.4-5.1) | 6.4 (2.9-10) | -3.3 (-1.3 to -8.1)<sup>2</sup> | -105 (-42 to -260)<sup>2</sup> |

<sup>1</sup>Due to acute COVID-19
<sup>2</sup>Negative sign indicates increase in cases with vaccination

**Figure 2.**
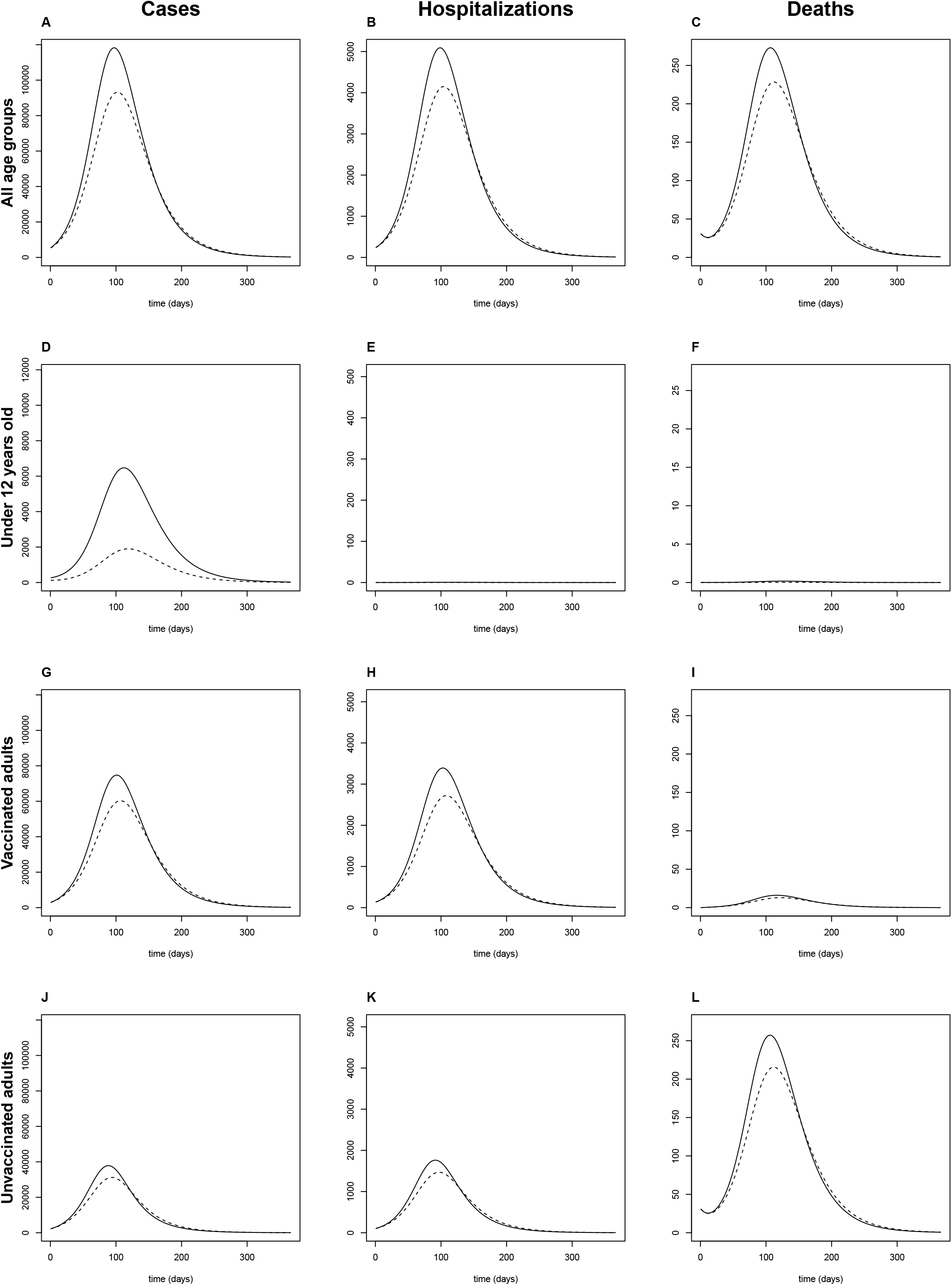
Projected wave of SARS-CoV-2 in Australia without (solid lines) and with (dashed lines) childhood vaccination (80% of children under 12 years of age). Using the SIR model, the epidemic curve was modelled over one year. Results for the population (all age groups) are shown in Panels **A-C**, and subdivided according to age and vaccine classes: children under 12 (Panels **D-F**), vaccinated adults (Panels **G-I**), and unvaccinated adults (Panels **J-L**). Model outcomes included daily incident cases (Panels **A, D, G**, and **J**), hospitalizations (Panels **B, E, H**, and **K**), and deaths (Panels **C, F, I**, and **L**). The expected waves of cases, hospitalizations, and deaths was reflected in the model, with modest reductions associated with childhood vaccination. Hospitalizations and deaths were infrequent in children under 12.

Circulation of SARS-CoV-2 strains with different transmissibility could alter model predictions. We therefore ran the model assuming *ℛ*_0_ = 2.79, corresponding to the alpha variant (Table S1). We also examined the scenario in which a higher proportion of adults was vaccinated (90%, Table S2).

Next, we performed sensitivity analyses to examine how model outputs varied with: (1) the proportion of children vaccinated; and (2) the intensity of concurrent public health measures. As the proportion of children vaccinated was varied between 0 and 1, an approximately linear relationship was observed in the reduction in cases, hospitalizations, and deaths across all age and vaccine classes (Figure S1). As the intensity of concurrent public health measures was varied, we found a non-linear relationship, with a local maximum in the reduction in cases at intermediate values of *θ* (Figure S2). A qualitatively similar pattern was observed for hospitalizations and deaths in all age and vaccine classes (Figure S3).

## Discussion

In light of the recent introduction of childhood vaccination against SARS-CoV-2, we forecast the effect on child and adult COVID-19 cases, hospitalizations, deaths, complications, and vaccine adverse events in two jurisdictions. Based on our mathematical model (Tables 2 and 3), several observations can be made for children 5-11 years of age: (1) high relative (percent) reduction in hospitalizations and deaths; (2) lower relative reduction in cases because of imperfect vaccine efficacy for prevention of transmission;^15^ (3) the absolute reduction in hospitalizations, deaths, and MIS-C was small, given the rarity of these events, even in unvaccinated children^16^; (4) cases of vaccine-associated myocarditis and anaphylaxis were few.^17^ For adults, modest herd immunity effects were observed, with relative reduction in hospitalizations and deaths on the order of 8-13%. Nonetheless, these correspond to non-trivial reductions in absolute numbers of hospitalizations (∼3700) and deaths (∼170), mostly among the unvaccinated. Cases of vaccine-associated myocarditis and anaphylaxis were predicted to increase, though case counts remained low.

In addition to projected population health impact, public acceptability will likely play an important role in the implementation of childhood vaccination against SARS-CoV-2. Seasonal influenza provides a benchmark vaccine-preventable respiratory virus against which SARS-CoV-2 public health impacts and vaccination can be compared. There were 349 SARS-CoV-2-related deaths in the first 19 months of the SARS-CoV-2 pandemic in the USA and 116 influenza deaths in the 2019-2020 season, respectively.^18,19^ Currently, many children in Australia and Alberta do not receive vaccination for seasonal influenza, suggesting that a substantial fraction of parents would be similarly reluctant to vaccinate their children against COVID-19.

Ethical considerations also arise around childhood vaccination. As shown in our model, by vaccinating children, they are benefiting the rest of the community. The argument in favour of this perspective is, however, difficult to prosecute because the people they will be protecting are those older individuals who refuse to be vaccinated or the small percentage of vaccinated people in whom the vaccine is ineffective. An additional ethical question is that of global social justice when administering vaccines to children in high-income settings while vulnerable elderly populations have limited access to vaccine in low-resource settings.

A higher relative impact of childhood vaccination (20-30% reduction in overall cases, hospitalizations, and deaths) was observed in models using the SARS-CoV-2 alpha variant with lower transmissibility (*ℛ*_0_ = 2.79, Table S1), and a higher baseline proportion of vaccinated adults (90%, Table S2). Moreover, childhood vaccination interacted synergistically with public health measures (*θ*) to produce a peak relative reduction in cases at intermediate values of *θ* (Figure S2). Taken together, these model predictions illustrate that under less intense epidemic conditions (when the effective reproduction rate, *ℛ*\*, is greater than, but close to, 1), modest reductions in the susceptible fraction (as occurs with childhood vaccination) can drive the epidemic toward extinction (*ℛ*\*<1), resulting in large relative reductions in the disease burden. On the other hand when *ℛ*\* is high, exponential growth continues with the modest reductions in transmission associated with childhood vaccination; therefore, the relative reduction in disease burden is minor. When *ℛ*\* is less than 1, the epidemic trends toward extinction with or without childhood vaccination; therefore, the relative reduction in disease burden is again minor. The implication for immunization programs is that childhood vaccination likely has the greatest potential for population-wide impact when coupled with other measures (e.g., social distancing, masking, adult vaccination).

Our modelling study has several limitations. The model was based on a deterministic compartmental SIR model; thus, stochastic effects were not considered.^10^ The system of 49 ODEs was parameterized with vital statistics from Australia or Alberta; therefore, extrapolation to other regions should be done with caution. On the other hand, the age structure and age-specific mortality rates were similar to other high-income, urbanized settings. Age-assortative mixing was incorporated into the model using social contact matrices; however, these were based on pre-pandemic surveys.^9^ Other investigators have demonstrated changes in contact rate with evolution of the pandemic, and have considered context-specific changes in contact rates (e.g. school closures).^20,21^ We used a composite ‘social distancing’ index (*θ*) to capture the combined effects of physical isolation, face masks, improved hand hygiene on the contact rate.^8^ This represents a simplification but is justified in the absence of data on the efficacy and uptake of various public health interventions in different age groups. The duration of infection prior to death or recovery was assumed to follow an exponential distribution. Recovered individuals in our model were considered immune (removed permanently from the susceptible pool); we did not incorporate waning immunity in the model. More complex mathematical formulations would be needed to reflect alternative distributions of the duration of infection and immunity. The co-circulation of several virus strains with different transmissibility was not incorporated into the model; however, for the simulation in Australia, we separately considered scenarios with circulating alpha variant (*ℛ*_0_ = 2.79) and delta variant (*ℛ*_0_ = 5.03). Vaccine efficacy may differ between strains of SARS-CoV-2, which could alter the model predictions. The impact of other vaccines (e.g., adenovirus-vectored vaccines) was not included in the model; however, mRNA vaccines are the primary vaccine product offered in Australia and Alberta currently.

In summary, our modelling results suggest that childhood vaccination yields modest benefits with minimal risk. Vaccination is predicted to result in substantial relative reductions in child morbidity and mortality, although the absolute reduction is small because these events are rare. Furthermore, the model predicts “altruistic” absolute reductions in adult cases, hospitalizations, and mortality, particularly among the unvaccinated, in whom the risk of these adverse outcomes is high.

## Data Availability

All data produced in the present study are available upon reasonable request to the authors

## Legends for Supplemental Materials

### Supplemental background information

Age-structured SIR model parameters

Characteristics of SARS-CoV-2 mRNA vaccine that inform model parameters

Multisystem inflammatory syndrome in children (MIS-C)

Vaccine adverse events

### Supplemental Results

Model predictions for a different SARS-CoV-2 strain (alpha variant)

Model predictions for higher vaccination rate among adults

Sensitivity analysis: proportion of children vaccinated

Sensitivity analysis: intensity of concurrent public health measures

**Table S1.** Simulation for Australia (*ℛ*_0_ = 2.79, 80% of adults vaccinated): Projected differences in cases, hospitalizations, deaths due to COVID-19, multisystem inflammatory syndrome in children (MIS-C) and vaccine adverse events associated with childhood vaccination.

|  | No childhood vaccination | Childhood vaccination<br>(80% coverage) | Absolute reduction | Relative reduction<br>(%) |
| --- | --- | --- | --- | --- |
| **Cases of COVID-19 (×1000)** |  |  |  |  |
| All age groups | 430 (88.1-7830) | 305 (72.5-6780) | 125 (15.6-1220) | 29 (13-37) |
| Under 12 years old | 15.5 (5.51-240) | 4.46 (1.26-115) | 11 (4.16-123) | 71 (52-78) |
| Vaccinated adults | 233 (28.4-5280) | 170 (24.6-4660) | 63 (3.74-738) | 27 (12-35) |
| Unvaccinated adults | 178 (50.4-2210) | 127 (43.6-1950) | 50.2 (6.72-382) | 28 (12-36) |
| **Hospitalizations<sup>1</sup>** |  |  |  |  |
| All age groups | 17800 (3590-337000) | 13000 (3130-297000) | 4750 (456-48300) | 27 (11-35) |
| Under 12 years old | 1.66 (0.592-25.8) | 0.479 (0.136-12.3) | 1.18 (0.447-13.3) | 71 (52-78) |
| Vaccinated adults | 10200 (1260-238000) | 7450 (1100-208000) | 2730 (162-33600) | 27 (11-35) |
| Unvaccinated adults | 7610 (2220-98700) | 5590 (1950-88900) | 2020 (274-15100) | 27 (11-35) |
| **Deaths<sup>1</sup>** |  |  |  |  |
| All age groups | 1630 (786-16600) | 1320 (745-14700) | 307 (41.7-2420) | 19 (5.2-30) |
| Under 12 years old | 0.569 (0.274-7.24) | 0.175 (0.135-1.51) | 0.393 (0.138-5.66) | 69 (50-82) |
| Vaccinated adults | 50 (6.31-1180) | 36.9 (5.49-1050) | 13 (0.797-174) | 26 (11-34) |
| Unvaccinated adults | 1580 (779-15500) | 1280 (738-13900) | 294 (40.7-2250) | 19 (5.1-30) |
| **MIS-C cases (0-19 years old)** | 4.89 (1.74-75.9) | 1.41 (0.399-36.3) | 3.48 (1.31-39) | 71 (52-78) |
| **Vaccine-related adverse events** |  |  |  |  |
| Myocarditis | 20 (9.0-120) | 38 (18-240) | -18 (-3.0 to -110)<sup>2</sup> | -93 (-15 to -570)<sup>2</sup> |
| Anaphylaxis | 22 (9.8-35) | 42 (19-68) | -20 (-8.1 to -50)<sup>2</sup> | -92 (-37 to -230)<sup>2</sup> |

<sup>1</sup>Due to acute COVID-19
<sup>2</sup>Negative sign indicates increase in cases with vaccination

**Table S2.** Simulation for Australia (*ℛ*_0_ = 5.08, 90% of adults vaccinated): Projected differences in cases, hospitalizations, deaths due to COVID-19, multisystem inflammatory syndrome in children (MIS-C) and vaccine adverse events associated with childhood vaccination.

|  | No childhood vaccination | Childhood vaccination<br>(80% coverage) | Absolute reduction | Relative reduction<br>(%) |
| --- | --- | --- | --- | --- |
| **Cases of COVID-19 (×1000)** |  |  |  |  |
| All age groups | 9510 (321-17200) | 7010 (150-16200) | 2510 (166-3140) | 26 (5.8-71) |
| Under 12 years old | 589 (29.6-971) | 154 (3.43-501) | 435 (26.5-535) | 74 (46-93) |
| Vaccinated adults | 7130 (167-14100) | 5440 (78.9-13500) | 1690 (76.7-2240) | 24 (3-70) |
| Unvaccinated adults | 1660 (109-1990) | 1320 (55.8-1890) | 345 (53.3-653) | 21 (5.1-69) |
| **Hospitalizations<sup>1</sup>** |  |  |  |  |
| All age groups | 398000 (12200-785000) | 305000 (6350-757000) | 93300 (5720-122000) | 23 (3.8-69) |
| Under 12 years old | 63.3 (3.18-104) | 16.6 (0.369-53.9) | 46.7 (2.85-57.4) | 74 (46-93) |
| Vaccinated adults | 323000 (7240-694000) | 243000 (3480-659000) | 79800 (3480-101000) | 25 (4-69) |
| Unvaccinated adults | 74800 (4570-101000) | 61400 (2470-98100) | 13400 (1780-26300) | 18 (2.6-67) |
| **Deaths<sup>1</sup>** |  |  |  |  |
| All age groups | 13900 (1150-20900) | 11100 (834-20200) | 2740 (314-4400) | 20 (3.6-60) |
| Under 12 years old | 17.7 (0.986-29.3) | 2.45 (0.174-5.78) | 15.2 (0.813-23.5) | 86 (80-93) |
| Vaccinated adults | 1620 (34.6-3700) | 1210 (17-3550) | 409 (16.7-528) | 25 (4.5-68) |
| Unvaccinated adults | 12200 (1120-17300) | 9930 (818-16700) | 2320 (302-4010) | 19 (3.3-59) |
| **MIS-C cases (0-19 years old)** | 186 (9.35-307) | 48.8 (1.09-158) | 137 (8.38-169) | 74 (46-93) |
| **Vaccine-related adverse events** |  |  |  |  |
| Myocarditis | 20 (9.0-120) | 38 (18-240) | -18 (-3.0 to -110)<sup>2</sup> | -93 (-15 to -570)<sup>2</sup> |
| Anaphylaxis | 22 (9.8-35) | 42 (19-68) | -20 (-8.1 to -50)<sup>2</sup> | -92 (-37 to -230)<sup>2</sup> |

<sup>1</sup>Due to acute COVID-19
<sup>2</sup>Negative sign indicates increase in cases with vaccination

**Figure S1.**
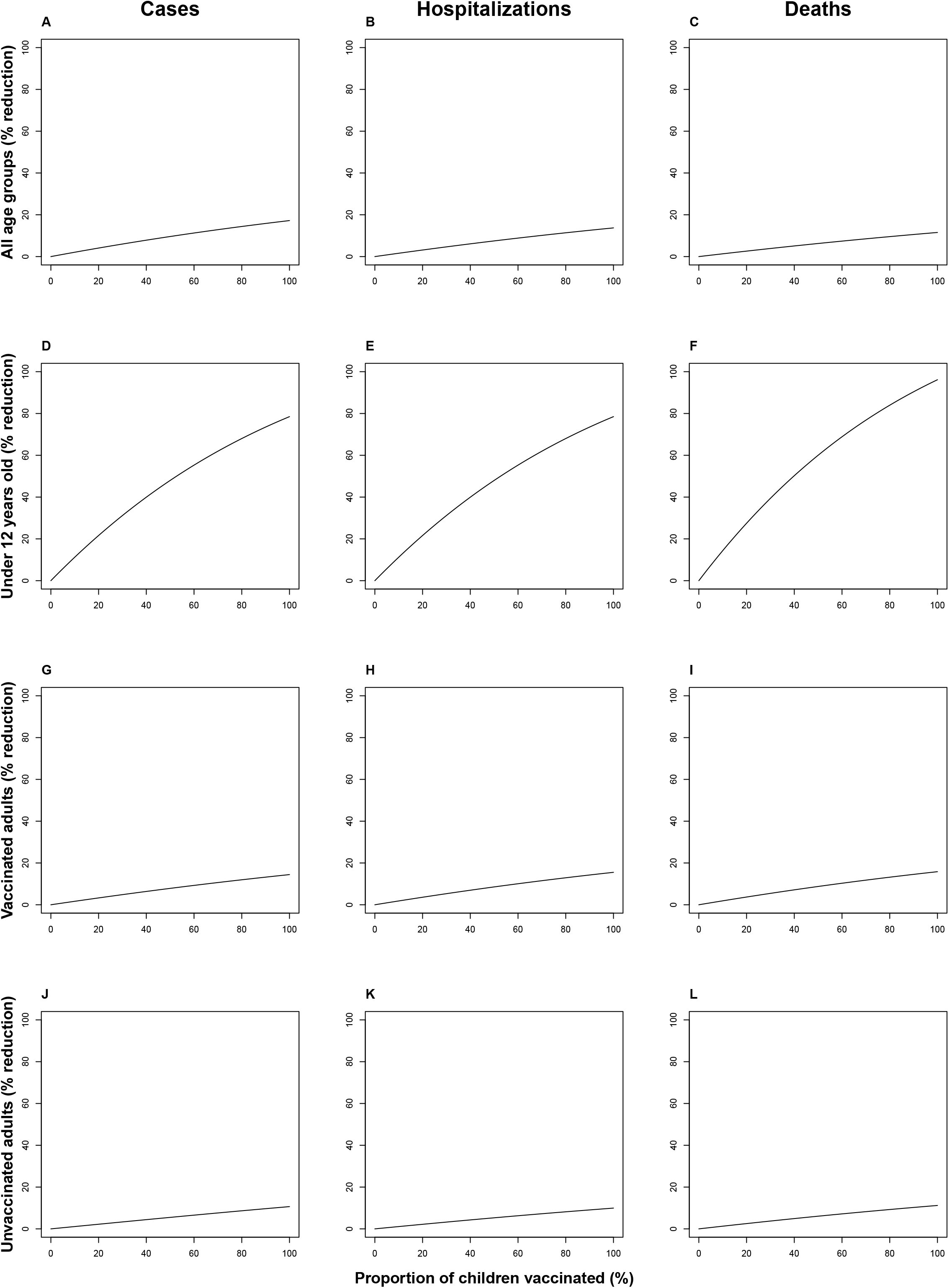
Sensitivity analysis of relative reduction in cases, hospitalizations and deaths, varying the proportion of children vaccinated. The relative (percent) reduction in daily incident cases (Panels A, D, G, and J), hospitalizations (Panels B, E, H, and K), and deaths (Panels C, F, I, and L) for all age groups (Panels A-C), children under 12 (Panels D-F), vaccinated adults (Panels G-I), and unvaccinated adults (Panels J-L) are shown with varying childhood vaccination rate. Of note, larger relative effects are seen in children (directly protected through vaccination) than adults (indirectly protected through increased herd immunity) over the range of vaccine uptake. The linear relationship predicts proportional reductions in cases, hospitalizations, and deaths with increasing vaccine uptake, with greatest relative reductions in hospitalizations and deaths in the under 12 age group (Panels E and F) and more modest relative reductions in other age and vaccine classes.

**Figure S2.**
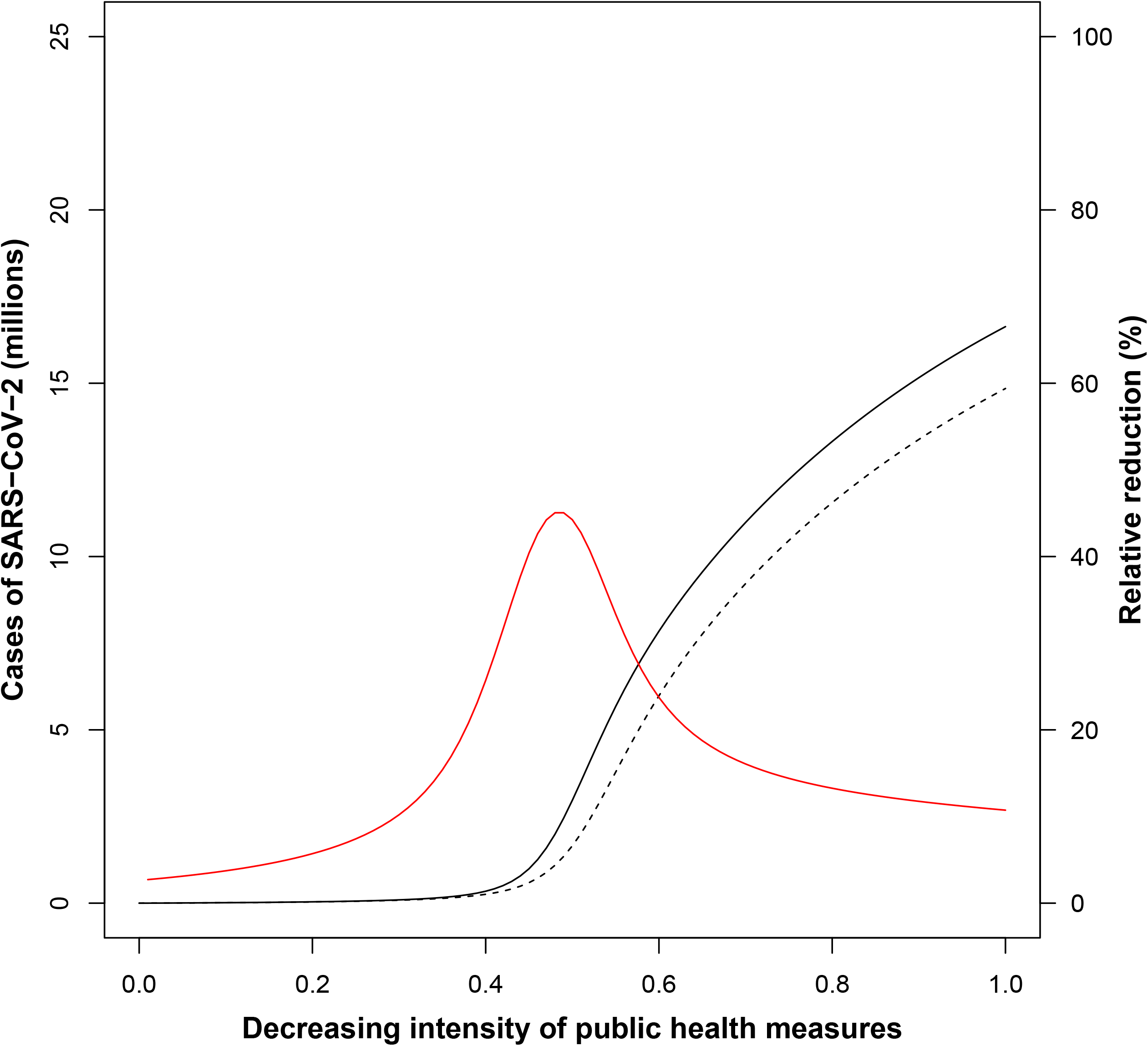
Sensitivity analysis: total cases (over 1 year of epidemic) vary with the intensity of public health prevention measures, as well as childhood vaccination. The solid black line indicates the scenario with no childhood vaccination, the dashed line indicates the scenario with 80% children vaccinated, and the solid red line indicates the relative (percent) reduction. The parameter theta (*θ*) was used to model changes in the contact rate with public health measures (e.g., social distancing, masking, business and service closures) that will likely continue to be necessary with the highly contagious delta variant (*ℛ*_0_ = 5.08). In the absence of any control measures (*θ* = 1), the SIR model predicted that a large fraction of the population (*N*_*T*_ = 25 million) would become infected over the course of the epidemic fourth wave, despite high vaccine coverage. The fraction of infected individuals decreased non-linearly with increasing public health measures. The effect of childhood vaccination was to qualitatively “shift” the curve toward the right. The relative reduction in cases with childhood vaccination yielded a curve with a local maximum around *θ* = 0.5. This interaction between vaccine coverage and intensity of public health measures can be explained as follows: with strict lockdown, transmission is low, the epidemic is extinguished, and is barely influenced by childhood vaccination; without control measures, transmission of highly infectious delta variant is not prevented by vaccinating children (a small fraction of the total population); but when moderate control measures are in place, reducing the effective reproduction number to near unity (*ℛ*\*≈1), the added benefit of childhood vaccination can “tip” the epidemic toward extinction, yielding a large relative reduction in total cases.

**Figure S3.**
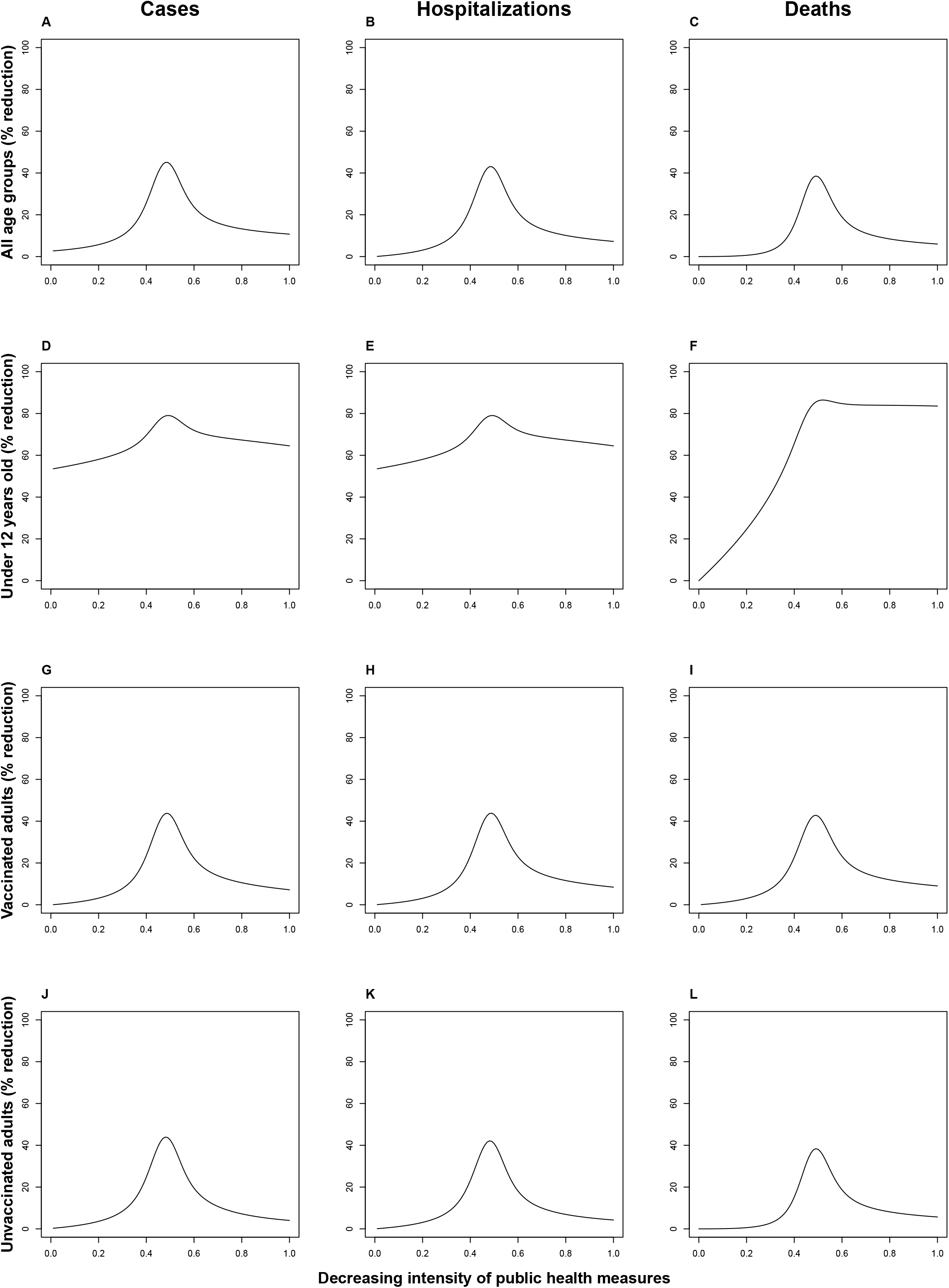
Sensitivity analysis of relative reduction in cases, hospitalizations and deaths, varying the intensity of public health measures. By varying theta (*θ*), a parameter used to model changes in the contact rate with public health measures, from 0 (complete lockdown) to 1 (perfect mixing), we estimated the relative reduction in incident cases (Panels A, D, G, and J), hospitalizations (Panels B, E, H, and K), and deaths (Panels C, F, I, and L) for all age groups (Panels A-C), children under 12 (Panels D-F), vaccinated adults (Panels G-I), and unvaccinated adults (Panels J-L). Model estimates for relative reduction in cases, hospitalizations, and deaths were sensitive to *θ*, reaching a local maximum around *θ* = 0.5. This suggests that childhood vaccination and public health measures interact, yielding maximal combined effectiveness at intermediate levels of social distancing.

### Supplemental background information

#### Age-structured SIR model parameters

Age-specific contact rates were quantified using published social contact matrices, specific for each geographic setting.^1^ In the referenced studies, these contact matrices were determined by survey methodology (number of people in each age stratum who engaged in a two-way conversation involving three or more words in the physical presence of the other person or direct physical contact) and statistical projections based on demographic information about the country.^1^ We used a series of transformations to account for the desired age group divisions and to satisfy the condition of reciprocity between age groups.^2^

Age-specific susceptibility to SARS-CoV-2 was incorporated into the model, based on the observation that, compared to a baseline risk in adults 15 to 64 years of age, children 0 to 14 years of age were less susceptible (odds ratio 0.34), and elderly individuals more than 65 years of age were more susceptible (odds ratio 1.47).^3^ Age variation in infectiousness was also included in the model, based on a study from Israel indicating that the relative infectiousness for children and youth younger than 20 years of age was 0.85, compared to adults.^4^ Other studies support the lower infectiousness of children.^5-7^

#### Characteristics of SARS-CoV-2 mRNA vaccine that inform model parameters

We reviewed the published literature to determine the best available data for vaccine efficacy. There are several possible clinically relevant outcomes of interest, including prevention of incident cases (through reduction in infectiousness and/or susceptibility to acquiring infection), hospitalizations, and deaths. We focused on the leading BNT162b2 mRNA vaccine (Comirnaty, Pfizer) since this vaccine has been most widely administered in North America and Australia and has the most detailed efficacy data. Where possible, we included data specific to the SARS-CoV-2 delta variant, since this is currently the dominant strain globally.

Vaccination has been shown to decrease both peak viral load and the duration of infection when breakthrough infection occurs.^8,9^ The efficacy of the vaccine in reducing infectiousness was directly studied among vaccinated and unvaccinated health workers in the UK.^10^ Household contacts had a reduced odds of acquiring SARS-CoV-2 from cases who had been vaccinated with the BNT162b2 mRNA vaccine compared to unvaccinated cases, although these data were collected during a phase of the epidemic when the alpha variant was circulating.^10^ In a retrospective cohort from Singapore, investigators were able to examine the vaccine efficacy against transmission of the delta variant.^11^ In addition, they were able to disentangle the effect of vaccination on susceptibility and infectiousness: the relative odds of household transmission was 0.33 if the contact was vaccinated compared to an unvaccinated contact (reduced susceptibility) and 0.73 if the index had been previously vaccinated compared to an unvaccinated index case (reduced infectiousness).^11^

SARS-CoV-2-related hospitalization is an important public health outcome, since epidemic waves threaten to overwhelm healthcare services. The vaccine efficacy in preventing hospitalization from SARS-CoV-2 was 86% (95%CI 82%–88%) in a recent analysis by the US CDC.^12^

With respect to mortality benefit of the vaccine, based on national surveillance data following a nationwide vaccination campaign in Israel, BNT162b2 mRNA vaccine was 96.7% effective at preventing death from SARS-CoV-2 (95% CI 96.0-97.3%).^13^

#### Multisystem inflammatory syndrome in children (MIS-C)

We included MIS-C as a distinct, serious, and occasionally fatal complication of COVID-19 in children.^14^ Incidence of MIS-C was estimated to be 316 persons per million SARS-CoV-2 infections in persons younger than 21 years.^15^ Deaths due to MIS-C are uncommon, and accounted for only 14% of decedents in a study of deaths due to COVID-19 among children and adolescents in the US.^16^

#### Vaccine adverse events

With respect to adverse reaction associated with COVID-19 mRNA vaccines, two distinct self-limited cardiac syndromes, myocarditis and pericarditis, have been reported.^17^ These adverse events have not been studied in children under 12 years of age. Therefore, we extrapolated the rate of these adverse events from available data in older children and young adults. Myocarditis occurred soon after immunization, in younger patients, mostly after the second vaccination.^17^ Pericarditis affected older patients later, after either the first or second dose.^17^ Because we were interested in the effects of childhood vaccination, we modelled the increase in myocarditis cases, since this was the childhood-specific severe adverse event.^18^ The incidence of myocarditis has been estimated at 10 to 63 per million doses with the highest incidence among males aged 12™17 years.^17,18^ No deaths have been reported for young adults who developed myocarditis after being given the mRNA vaccines, despite 1,226 reports of myocarditis reported in the US. However, a single death due to myocarditis has recently been reported from New Zealand.^19^ Because of the apparent rarity of this event, deaths from vaccine-associated myocarditis were not counted in our model.

The US CDC detected 21 cases of anaphylaxis following 1.9 million doses of the Pfizer-BioNTech mRNA SARS-CoV-2 vaccine (11.1 cases per million doses).^20^ Most (71%) cases occurred within 15 minutes of vaccination and there were no fatalities.^20^ We included the predicted number of cases of anaphylaxis in our model; however, deaths from anaphylaxis were not counted.

## Supplemental Results

### Model predictions for a different SARS-CoV-2 strain (alpha variant)

Because of variations in transmissibility, co-circulation or change in the dominant circulating strain could affect model predictions. In addition to the delta variant (*ℛ*_0_ = 5.08) presented in the main manuscript, we modelled the course of the epidemic with the alpha variant (*ℛ*_0_ = 2.79). The results over one year are shown in Table S1.

### Model predictions for higher vaccination rate among adults

As of 28 Nov, 2021, 86.7% of Australians over the age of 16 years had received two doses of vaccine.^21^ We therefore considered a separate case in which the rate of adult vaccination was 90% (versus 80% in Table 2, main manuscript). The results over one year are shown in Table S2.

### Sensitivity analysis: proportion of children vaccinated

Next, we examined the effect of varying the proportion of children vaccinated on the model outputs. A linear relationship was observed in absolute reduction (data not shown) and relative reduction in cases, hospitalizations, and deaths in all age and vaccine classes (Figure S1). Conveniently, this linear relationship would allow the predicted effect of less complete vaccine coverage to be calculated in a straightforward manner.

### Sensitivity analysis: intensity of concurrent public health measures

In a sensitivity analysis of the effect of childhood vaccination, varying the intensity of public health measures (*θ* in the SIR model), we found a non-linear relationship with a local maximum in relative reduction of SARS-CoV-2 cases at intermediate values of *θ* (Figure S2). A qualitatively similar pattern was observed for the number of hospitalizations and the number of deaths (data not shown). A similar pattern was seen for other model outcomes (hospitalizations and deaths) in all age and vaccine classes (Figure S3). This analysis suggests an interaction between childhood vaccination and public health measures, with highest effect of childhood vaccination when some public health measures remain in place.

